# Brief Communications: Phenome-wide Association Study of Cognitive Function in Multiethnic Asian population

**DOI:** 10.1101/2025.06.18.25329738

**Authors:** Theresia Mina, Nilanjana Sadhu, Dorrain Yanwen Low, Pritesh R Jain, Xiaoyan Wang, Hong Kiat Ng, Tim van Der Es, Keane Lim, The HELIOS Study team, Paul Elliott, Elio Riboli, Joanne Ngeow, Lee Eng Sing, Sabrina Wong Kay Wye, Kok Pin Ng, Marie Loh, Jimmy Lee, John Chambers, Max Lam

## Abstract

Cognition is one of the most widely used measure of brain health. Poor cognitive function reflects both the progression of neurodegenerative disorders and the decline of cardiometabolic conditions. Moreover, the genetic determinants of cognitive function are pleiotropic with various aspects of physical health and mortality. To date, little research representing the diverse Asian populations has been done. To understand the epidemiological role of cognition and its biological underpinnings in this population, we performed PheWAS on 542 unique research variables collected in the HELIOS Study (Singapore), with a dataset comprising 8207 individuals (Chinese: Indian: Malay= 69:18:13) aged 30-84 years in whom ***‘g’,*** general factor for cognition, was derived. We used ***‘g’*** as a predictor and adjusted for age, sex, ethnicity, the interaction of age and sex and genetically determined ancestries. We then performed 2-sample Mendelian Randomization and downstream sensitivity analyses using the GWAS summary statistics from the UK Biobank to ascertain causality. We found that ***‘g’*** was associated with a broad range of phenotypes (Bonferroni p<8.1x10^-5^, 148 [27.3%] phenotypes), ranging from greater familiarity with digital devices (β=0.21, p=5.5x10^-138^), to smaller waist-hip-ratio (β =-0.06, p=2.5x10^-15^) and lower carbohydrate intake (β=-0.07, p=3.5x10^-11^). After further hierarchical clustering to reduce redundancies and ensure representativeness across domains, we shortlisted 23 (4.2%) representative phenotypes. The Inverse Variance Weighted 2-sample MR demonstrated causal linkage of genetically predicted ***‘g’*** with 14 demographic and physiological phenotypes, including less body fat percentage, lower alcohol consumptions, higher polyunsaturated/monounsaturated fatty acids (PUFA/MUFA) consumptions, household income and total year of education, with significant horizonal pleiotropy. From the initial 147 SNPs in the instrument-exposure ***‘g’***, we selected 134 SNPs which uniquely predicts ‘***g***’ and performed further annotations. The SNPs are linked with genes involved in neural development, neurotransmission, neuroprotection, and chromatin modification and transcriptional regulation. Our study demonstrates broad implications of cognitive function in lifestyle choices and cardiometabolic health indices as potential targets for early health maintenance at population levels. Furthermore, the HELIOS Study addresses the diversity gap in the cognitive genetic research and presents an opportunity to improve the cardiometabolic and cognitive health in this multiethnic Asian populations.

## Introduction

The global population growth has stagnated, leading to the rapid ageing^1^. As age represents the greatest risk factor for the onset of dementia, it posits tremendous social and economic impacts, with estimated costs surpassing 2.8 trillion by 2030.^2^ Cognitive function is the hallmark of healthy ageing, as it is protective against future risk of dementia owing to neurodegenerative diseases such as Alzheimer’s Disease (AD)^3^. Beyond neurological health, cognitive performance has been proposed as an endophenotype of overall ‘system integrity’, with broad effect on sociodemographic outcomes and physical wellbeing.^4^ Moreover, cognitive function is genetically correlated with various psychopathologies, including autism spectrum disorders, schizophrenia, and Major Depressive Disorder (MDD).^5^ Understanding the consequences of cognitive health and its biological underpinnings could help maintaining human potentials and wellbeing, and are therefore of significant public health importance.

Phenome-Wide Association Study (PheWAS) allow the hypothesis-free identifications of cognitive health outcomes, given the potentially wide ranges of health consequences. When applied to epidemiological cohort data, PheWAS allows comparisons of health parameters across biological systems objectively, as often performed in large biobanks such as the UK Biobank (UKB). The global burden of dementia has also geographically shifted towards Asia-Pacific, with the 2025 World Alzheimer Report estimated that Asian populations contribute towards ∼40% incident dementia cases globally.^6^ However, European-centric PheWAS might have limited applications in Asian populations; the heterogeneity in health outcomes could be driven by ancestry, as observed in the varying impact of *APOE*ε4 frequency on AD^7^, or by lifestyle risk factors, as in the variation of dietary risk factors for cardiovascular diseases across world regions.^8^ Therefore, to better understand the consequences of impaired cognitive health in Asia-Pacific and globally, we conducted PheWAS of cognitive function as exposure variables using epidemiological data from ∼10,000 participants in the Health for Life in Singapore (HELIOS) Study, a multi-ethnic Asian population cohort study in Singapore^9^, and followed-up with Two-Sample MR of cognitive function as instrument-exposure on the candidate outcome phenotypes using summary statistics from independent populations.

## Results

### Cognitive function confers impact on wide ranges of phenotypic domains in multi-ethnic Asian populations

We reviewed a catalogue of 3190 variables from the HELIOS study and after aggregations and derivation of summary variables, we shortlisted a total of 542 variables suitable for downstream PheWAS (**S Fig 1**). The socio-demographic assessment is the largest variable contributors (265; 48%), whereas hand grip is represented by only 2 variables. Up to 9,296 Chinese, Indian and Malay participants completed cognitive function test to enable the calculation of ‘***g***’, and a subset of 8,207 had successful blood draw leading to genetic data (**S Fig 1**). In the 8,207 dataset, participants were 58.5% female, 52.05 (11.43) years, with 69:18:13 Chinese: Indian: Malay ethnicity ratio (**S Table 1**).

**Figure 1.**
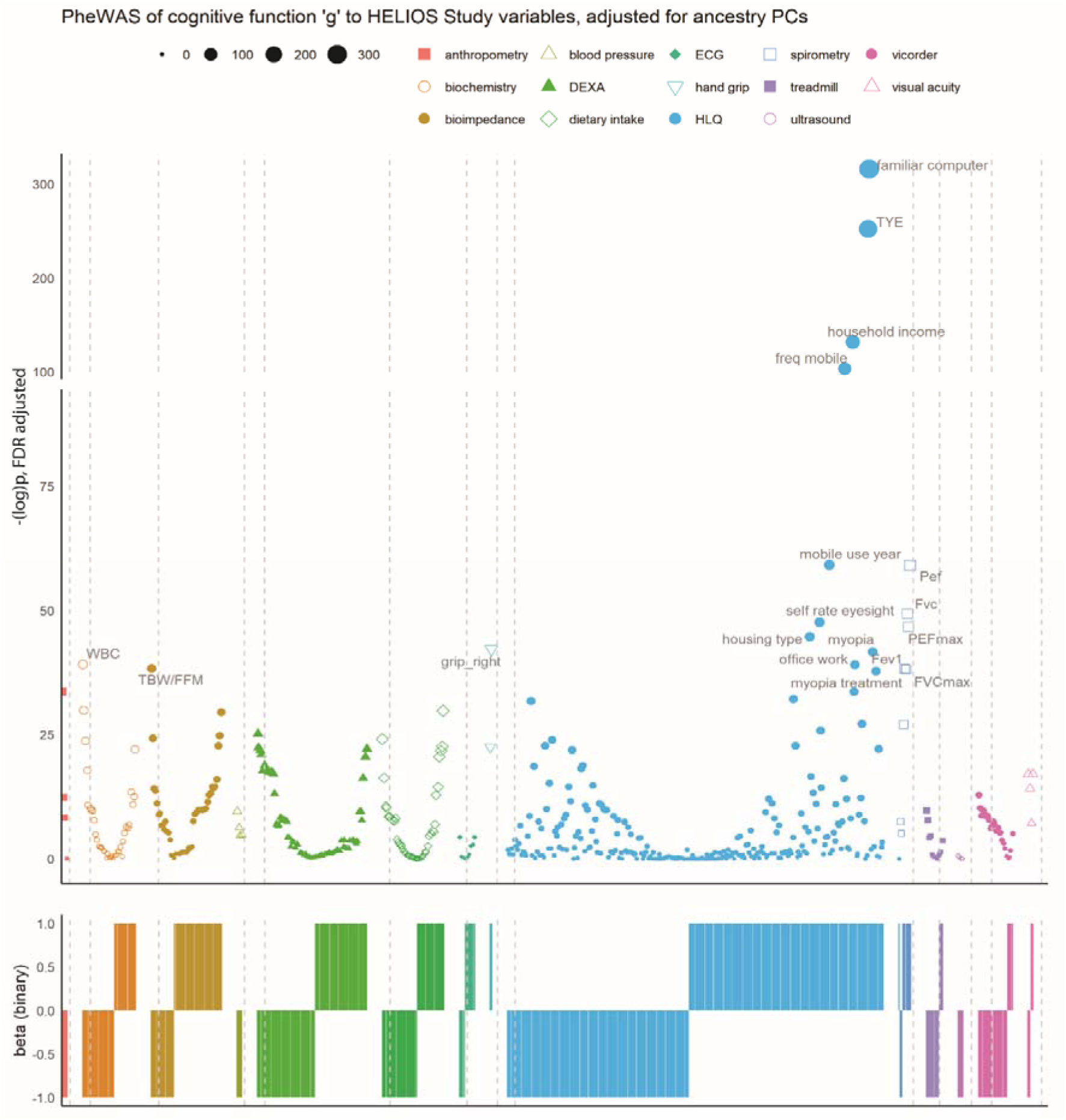
Phenome-wide Association Studies (PheWAS) of general cognitive function ‘*g*’ in multiethnic Asian populations. PheWAS was conducted with ‘*g*’ as exposure variable and 542 research variables as outcome variables. The research variables were grouped according to the HELIOS Study assessments (x-axis), in alphabetical order of assessments. Colour and shape represent different assessment domains, separated by grey dotted lines. Circle size represents the magnitude of -log(P). All continuous variables were z normalised. All regressions were adjusted for sex, age, ethnicity, the interaction of sex and age, age-squared, sex and age- squared, and the first 5 genetic PCs. The variable coding was such that higher scores indicate greater values of outcome variables, unless otherwise indicated. To illustrate the direction of the associations, β was converted to binary outcome. *DEXA*: Dual X-Ray Absorptiometry; *ECG*: electrocardiogram; *TYE*: total year of education; *freq mobile*: frequency of mobile phone usage/week; *FEV1*: Forced Expiratory Volume; *FVC*: Forced Vital Capacity; *HLQ*: health and lifestyle questionnaire; *PEF*: Peak Expiratory Flow; *WBC*: white blood cell; *WHR*: waist-hip ratio.

**Table 1.** GWAS summary statistics of instrument-outcome phenotypes predicted by general cognitive function ‘*g*’. The summary statistics were restricted to Europeans, with larger sample size and later publication year prioritised. Abbreviations: *FEV1*: Forced Expiratory Volume; *FFM*: fat-free mass; *FVC*: Forced Vital Capacity; *MUFA*: mono-unsaturated fatty acids; *PEF*: Peak Expiratory Flow; *PUFA*: poly-unsaturated fatty acids; *TBW*: total body water; *vFMI*: DEXA-based visceral fat mass index; *WBC*: white blood cell.

| Domain | HELIOS Variable | OpenGWAS (restrict to European) | Consortium | year | OpenGWAS ID | n |
| --- | --- | --- | --- | --- | --- | --- |
| Socio-demographics x 8 | total year of education | Years of schooling | SSGAC | 2018 | ieu-a-1239 | 766,345 |
|  | Eyesight, myopia, wear glasses | Reason for glasses/contact lenses: For short-sightedness | MRC-IEU | 2018 | ukb-b-6353 | Case: 37,362<br>Control: 423,174 |
|  | Familiar with computer | Time spent using computer | MRC-IEU | 2018 | ukb-b-4522 | 360,895 |
|  | Frequency and year using mobile phone | Length of mobile phone use | MRC-IEU | 2018 | ukb-b-4094 | 456,972 |
|  | Household income | Average total household income before tax | MRC-IEU | 2018 | ukb-b-7408 | 397,751 |
|  | Housing type | Own or rent accommodation lived in: Rent - from local authority, local council, housing association | MRC-IEU | 2018 | ukb-d-680_3 | Case: 18,968<br>Control: 337,372 |
|  | Office work | Job involves heavy manual or physical work | MRC-IEU | 2018 | ukb-b-2002 | 263,615 |
|  | Own car | Transport type for commuting to job workplace: Public transport | MRC-IEU | 2018 | ukb-b-17155 | 245,364 |
| Body composition x 5 | Height | Height | Yengo et al | 2018 | ebi-a-GCST90029008 | 673,878 |
|  | Percent body fat | Body fat percentage | MRC-IEU | 2018 | ukb-b-8909 | 454,633 |
|  | TBW/ FFM | Whole body water mass; | MRC-IEU | 2018 | ukb-b-14540; | 454,888 |
|  |  | Whole body fat-free mass |  |  | ukb-b-13354 | 454,850 |
|  | vFMI | Visceral Adipose Tissue (VAT) | UKB | 2019 | GCST008744 | 325,153 |
| Biochemistry x 3 | Monocytes | Monocyte count | Chen MH et al | 2020 | ebi-a-GCST90002340 | 521,594 |
|  | neutrophil | neutrophil cell count | Chen MH et al | 2020 | ebi-a-GCST90002351 | 519,288 |
|  | WBC | White blood cell count | Chen MH et al | 2020 | ebi-a-GCST90002374 | 562,243 |
| Dietary habit x 3 | Alcohol consumption | Alcohol intake frequency | MRC-IEU | 2018 | ukb-b-5779 | 462,346 |
|  | Carbohydrates | Carbohydrate | MRC-IEU | 2018 | ukb-b-7244 | 64,979 |
|  | MUFA | MUFA/ PUFA | Richardson TG et al | 2022 | ebi-a-GCST90092940 | 115,006 |
| Spirometry x 3 | FEV1 | FEV in 1-second | MRC-IEU | 2018 | ukb-b-19657 | 421,986 |
|  | FVC, FVCmax | FVC, best measure | MRC-IEU | 2018 | ukb-b-14713 | 345,665 |
|  | PEF, PEFmax | PEF | MRC-IEU | 2018 | ukb-b-12019 | 421,986 |
|  | Grip strength, right | Hand grip strength (right) | MRC-IEU | 2018 | ukb-b-10215 | 461,089 |

**##Figure 1** highlighted 148 out of 542 (27.3%) research phenotypes associated with ‘***g***’ across assessment domains with Bonferroni p<8.1x10^-5^. Greater ‘***g***’ (reflecting higher cognitive function) was associated with broad range of phenotypes, including greater familiarity with digital devices such as computer (β=0.21, p=5.5x10^-138^**, S Table 2**), smaller waist-hip-ratio (β =-0.06, p=2.5x10^-15^**, S Table 2**) and lower carbohydrate intake (β=-0.07, p=3.5x10^-11^**, S Table 2**). The socio-demographic domain contains the top 5 associations (familiarity with digital devices, and subsequently total year of education, household income, frequency of mobile phone usage, and years of using mobile phone, β range=|0.10|; |0.19|, p range=2.0x10^-26^; 2.9x10^-110^**, S Table 2**), followed by several measures of lung function (β range=|0.04|; |0.09|, p range=0.0006; 5.3x10^-21^**, S Table 2**). We performed further sensitivity PheWAS by substituting ‘***g***’ with reaction time as alternative exposure of cognitive function, because reaction time is reported to be orthogonal to ‘***g***’^5^. The analysis with reaction time as the main exposure variable also remained broadly similar with the main findings (**S Fig 2, S Table 3**).

**Figure 2.**
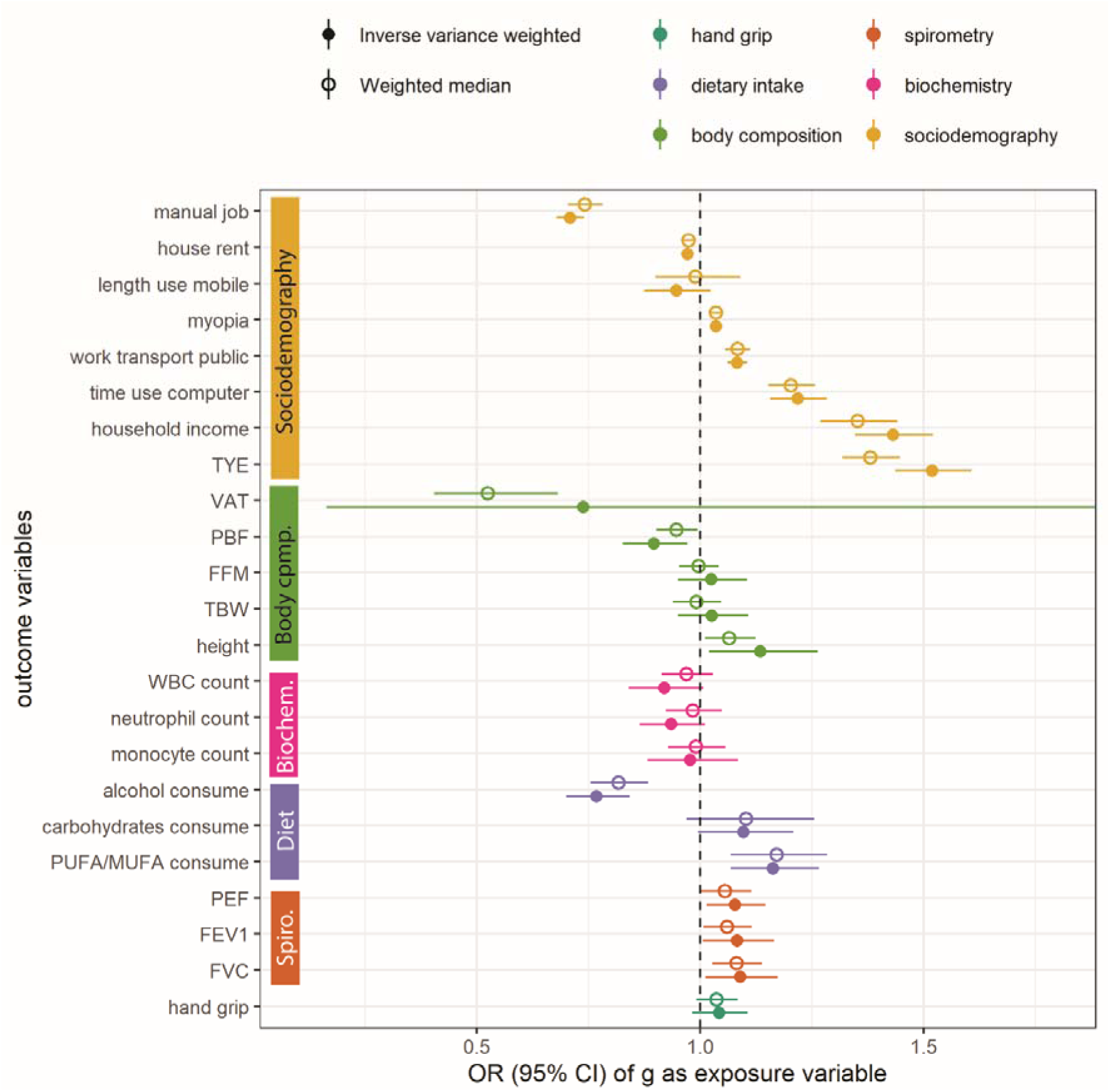
Two-sample Mendelian Randomisation (MR) of general cognitive function ‘*g*’ following PheWAS. The ‘*g*’ was instrument-exposure. *FEV1*: Forced Expiratory Volume; *FFM*: fat-free mass; *FVC*: Forced Vital Capacity; *MUFA*: mono-unsaturated fatty acids; *PBF*: Percent Body Water; *PEF*: Peak Expiratory Flow; *PUFA*: poly-unsaturated fatty acids; *TBW*: total body water; *TYE*: total year of education; *VAT*: DEXA- based visceral fat mass; *WBC*: white blood cell.

Having identified enrichment of phenotypes associated with ‘***g***’ at p<10^-50^ in the socio- demographic and spirometry domains, we subsequently conducted hierarchical clustering on the intra-correlations of phenotypes across other assessment domains to further probe phenotypes with p at the marginal regions of **Figure 1**, arbitrarily defined as between -log(p)=30 to 35 (9.3x10^-14^ to 6.3x10^-16^), and attempted to shortlist at least 1 representative variable per domain (**S Fig 3**). Overall, we shortlisted 23 (4.2%) phenotypes predicted by general cognitive function **‘*g*’ (Table 1).** No further clustering was performed on blood pressure, ECG, carotid ultrasound and visual acuity assessments due to the small number of variables. Notably, greater ***‘g’*** was associated with reduced blood pressure and heart rate and ventricular rate (β range= -0.02; -0.04, p range=0.01;7.35x10^-5^, **S Table 2**), though they did not achieve marginal p-value cut-off. The associations of ***‘g’*** with carotid ultrasound were not significant (p>0.701**, S Table 2**). None of the ‘***g***’ associations with the vicorder (arterial stiffness) and treadmill phenotypes achieved the Bonferroni correction threshold p∼9.3x10^-14^ (**S Table 2, S Fig 3**). No associations of ‘***g***’ with visual acuity assessments (β range =|0.01|; |0.05|, p range=0.0008; 4.0x10^-8^) were found to cross the multiple testing correction. Nonetheless, ophthalmological health is represented by the associations of ‘***g***’ with self-rated eyesight and myopia (β(p)= 0.09(2.0x10^-21^) and 0.23(8.4x10^-19^), respectively, **Figure 1, S Table 2**).

### Cognitive function is causally linked with a wide, multi-domain array of health phenotypes

We next curated the GWAS summary statistics of the representative instrument-outcome phenotypes to facilitate the Two-Sample Mendelian Randomisation (MR) to demonstrate causality linking ‘***g***’ and the 23 shortlisted phenotypes. Given the limited availability of open-sourced GWAS summary statistics derived from Asian populations, we focused on the UK Biobank-generated summary statistics derived from European individuals in the OpenGWAS catalog (https://gwas.mrcieu.ac.uk/). We included summary statistics representing the 23 epidemiological phenotypes (**Table 1**). No summary statistics represented “*office work*”, hence we selected “*job involves heavy manual or physical work*” (ukb-b-2002, **Table 1**). We selected binary status of renting (ukb-d-680_3, **Table 1**), because of the close to 90% of house ownership in Singapore^10^, compared with 63% in England.^11^ We derived the GWAS summary statistics for ‘***g***’ from the meta-analysis of Cognitive Genomic Consortium, Social Science Genetic Association Consortium, and UKB^12^. The F-Statistics for all instruments were >10 (**S Table 4**).

After clumping and harmonisation, the number of Single Nucleotide Polymorphisms (SNPs) ranged from 145 to 148 across all instrument-outcome pairs, except for Visceral Adipose Tissue (VAT), for which only 8 SNPs remained for the downstream MR. Following Inverse Variance Weighted (IVW) and weighted median MR, greater ‘***g***’ is causally linked with 14 phenotypes, including greater computer usage, household income and total year of education, higher myopia risk, likelihood of using public transport, greater height, PUFA/MUFA consumptions, and lung functions—Peal expiratory flow (PEF), Forced expiratory volume (FEV1) and Forced vital capacity (FVC) (**Figure 2, S Table 5**). Greater ‘***g***’ is also causally linked with lower likelihood of renting house and having a manual job, lower percentage body of fat, and lower alcohol consumptions (**Figure 2, S Table 5**). Greater ***‘g’*** was neither causally associated with higher fat-free mass or stronger grip strength, nor with less white blood cell counts (**Figure 2, S Table 5**).

Among the MR-Egger intercepts (**S Table 6**), only the analysis with myopia as the outcome showed a p<0.05, suggesting horizontal pleiotropy. The leave-one-out MR pleiotropy (**S Fig 4**) and the Single SNP MR (**S Fig 5**) implied horizontal pleiotropy in some instrument-outcomes, such as height, PUFA/MUFA consumption, household income and total year of education. The Q-statistics also demonstrate significant heterogeneity across instruments (**S Table 7**). From the initial 147 SNPs in the instrument-exposure ‘***g***’, there were 1062 SNP-phenotype combinations available in the GWAS catalogue, including pleiotropic SNPs (**S Fig 6, S Table 8**). After removing pleiotropic SNPs associated with non-cognition phenotypes and/or other psychiatric conditions, 134 unique SNPs were available in the instrument-exposure. The repeat MR without pleiotropic SNPs demonstrate robust associations between genetically predicted ‘***g***’ and the 11 physiological and sociodemographic phenotypes (**S Fig 7, S Table 9**), similar to **Figure 2**.

In the reverse MR where ‘***g***’ was the instrument-outcome, less likelihood of manual job, higher household income, and total year of education, lower VAT, percentage body fat, greater height and total body water, fat-free mass, greater lung function and grip strength were all causally linked with higher ‘***g***’ (**S Fig 8, S Table 10**). However, unlike earlier causal linkage between ‘***g***’ and ophthalmological health and dietary intake, higher ‘***g***’ was not predicted by less alcohol consumptions, higher myopia risk and PUFA/MUFA consumptions (**S Fig 8, S Table 10**).

Of the 134 non-pleiotropic SNPs predicting ‘***g***’, over 40% are located in the intronic regions (see Consequences, **S Fig 9, S Table 11)**, and 28% fall within protein-coding regions (see Biotype, **S Fig 9, S Table 11)**. Notably, most SNP-allele combinations (96 out of 348, 38.7%) had no affiliated candidate genes. As expected, candidate genes are over-expressed in brain sub-tissues (**S Fig 10**), and are generally involved in the modulation of chemical synaptic transmission, signalling and developmental cell growth (**S Fig 11**). There was 1 gene linked with 5 SNPs (*KRT18P1*), 1 with 4 SNPs (*GATAD2B*), and another 77 (22.1%) candidate genes represented by at least 2 SNPs. *GATAD2B* is vital for chromatin modification and transcriptional regulation, associated with intellectual disability and autism spectrum disorders.^13–15^ Other candidate genes include: *AUTS2*, which is implicated in numerous neurological disorders,^16^ *CAMKV*^17^ and *RBL2*^18^, which are important for neural development. Finally, we did not observe significant difference in the minor allele frequency of SNPs predicting ‘***g***’ across ancestries (P_ANOVA_=0.243, **S Fig 12, S Table 12**).

## Discussion

We demonstrated phenome-wide associations of cognitive function with a wide range of health parameters spanning across phenotypic domains in a multiethnic Asian population. Using two-sample MR analysis in an independent European population, we highlighted causal linkage of higher cognitive functions with higher education attainment, household income, likelihood of home ownership, better lifestyle choices and cardiovascular parameters including greater lung functions and less total body fat. Our findings reveal the importance and potentials of cognitive health maintenance in individual wellbeing, ranging from better socio-economic profile to more favourable physiological outcomes. As genetic variation was randomly assigned at birth, our findings elucidate the biological relationship linking cognitive health with human potentials and health traits not confounded by individual environmental constructs.

We show causal linkage of higher cognition predicting higher PUFA/MUFA consumptions and lower percentage of body fat, but not visceral fat. This is perhaps unsurprising, given that subcutaneous fat, which largely contributes to total body fat, has distinct genetic makeup and functionalities compared with visceral fat^19–21^. We also did not observe reverse causality linking carbohydrate or PUFA/MUFA consumptions with ‘***g***’ found in paediatric populations,^22^ implying potential temporality in the impact of diet on cognitive function. Our tissue over-expression analysis supports plausible hypothalamic role of appetite and satiety regulation,^23^ the role of appetite hormones on regulating memory,^24^ and hypothalamic regulation of lipogenesis and lipolysis.^23^ From evolutionary perspective, spatial memory bias towards high-calorie foods^25^ and increased body fat deposition provide survival advantage. However, selective pressure for increased fat deposition has reduced over time with greater dietary flexibility and technological inventions to store excess energy resources outside of the body^23^. Spatial food memory bias might be also maladaptive in the contemporary obesogenic food environment^26–28^ Recent meta-analysis revealed that behavioural interventions, including cognitive behavioural therapy, could promote a more sustainable, long-term weight loss compared with pharmacological interventions.^29^ Our study provides further motivations to incorporate cognitive health maintenance as part of the public health strategies to combat excess adiposity in Asia-Pacific and globally.

Our findings also have several implications on neurological and psychological wellbeing at population levels. Cognitive function is closely linked with (but not identical to) cognitive resilience; higher cognitive resilience could compensate against age-related neurodegenerative brain changes via increased functional efficiency of brain networks or utilization of alternative networks to compensate for pathological disruption of pre-existing networks,^30,31^ thereby potentially delaying neurodegenerative onsets.^32^ Furthermore, the causal associations identified between cognitive function and multiple health measures in this study, together with previously reported Mendelian Randomization evidence demonstrating a protective effect of cognitive function against Alzheimer’s disease risk,^33^ underscore the importance of maintaining cognitive health as a preventive strategy against age-related neurodegeneration. On the other hand, despite the genetic correlations of MDD with reaction time^34^ and the pleiotropy of cognition with other psychopathologies^5^, we only identified cross-sectional associations between higher ‘***g***’ and lower levels of depression and anxiety symptoms, which did not achieve the marginal p-value cut-off for the follow-up MR analysis. Recent PheWAS revealed immaterial associations of polygenic risk score (PRS) for depression with cognitive ability.^35^ Different cognitive domain could also confer opposite effects on MDD, leading to null hypothesis.^5^ Nevertheless, even in the absence of causal linkage between cognition and depression, a meta-analysis demonstrated the long-term efficacy of cognitive behavioural therapy as treatment for depression, compared with other psychotherapies and pharmacotherapies.^29^

Importantly, our study also highlights the impact of cognition on social outcomes. A recent GWAS on income found genetic correlations between cognitive performance and income that became nonsignificant after adjusting for educational attainment.^36^ This suggests educational attainment largely mediates the causal impact of ‘***g***’ on social outcomes. This provides further motivations to use polygenic risk scores to understand the biological mechanisms on how genetic variants influence cognitive function and education attainment, and how it is modulated by environmental risk factors. We emphasise that cognitive function is shaped by a complex interplay of genetic and environmental factors, highlighting its potential modifiability over time.

Our investigation has some limitations. MR inferences are limited in temporality and assume linear, lifetime estimates of risk.^37^ We did not evaluate incident disease outcomes as this would require linkage with health records, but the initial linkage exercise of the first 10,000 participant data with the corresponding healthcare records, in close collaborations with the Singapore’s Ministry of Health, was sucesful^9^. As discussed previously,^38^ we acknowledge that the computerised nature of our cognitive test might have disadvantaged those with less digital proficiency. Individuals who were excluded from the analysis were also more likely to be less metabolically healthy, hence the reported epidemiological associations were likely to underestimate the potential cognitive health burden. We also could not ascertain the European-based Two-sample MR findings in Asian populations using One-sample MR using the HELIOS study data. At the point of the writing, the study is completing data collection up to 100,000 participants, with only preliminary GWAS performed on the first 10,000 participants for few phenotypes.^9^ However, the similarity of our epidemiological findings and Two-sample MR, coupled with the similar minor allele frequency of relevant SNPs predicting ‘***g***’ across ancestries, imply that findings should be generally applicable to Asian populations. As the Asian summary statistics were unavailable, we could not prevent participant overlaps to reduce bias between the UK Biobank and COGENT^12^, but we did perform multiple sensitivity analyses, including removing SNPs predicting alternative phenotypes. Further, in our previous MR exercise we demonstrated that the potential bias due to sample overlap between the UK Biobank and COGENT was minimal.^38^

Our study has several strengths. First, unlike prior MR studies investigating the impact of cognitive health on predefined physical and mental health traits,^39^ our MR was guided by a PheWAS approach, which is not constrained by *a priori* hypotheses and allows full utilisation of the available phenotypic information. Second, although the pleiotropy of cognitive performance with physical health is established,^39^ our sensitivity analyses removing pleiotropic SNPs demonstrated the robustness of causality, enabling the identification of candidate genes and biological pathways enriched in the brain and central nervous systems. Third, compared with the other large East Asian cohort studies such as China Kadoorie Biobank,^40^ BioBank Japan,^41^ Taiwan Biobank,^42^ and KoGES,^43^ our study is the first multi-ethnic Asian PheWAS exercise inclusive of Southeast and South Asian individuals. Fourth, the epidemiological and Two-sample MR findings were consistent, providing future basis for the construction of polygenic risk scores of cognitive function. The HELIOS Study participants were also recruited from the community, and are representative of the Singapore populations.^9,44^ Further, in the context of cognitive health, the cohort included participants aged 30 and above, who are relatively younger and of working age groups, and are therefore less likely to be confounded by underlying cognitive decline and reduce potential overestimations of the associations of ‘***g***’ with phenotypes. Finally, the study protocol, including cognitive test and all other physiological assessments, was also modelled after the UK Biobank, ensuring international operability.^9^

## Conclusion

This investigations demonstrates broad implications of cognitive function in multiethnic Asian populations. The HELIOS Study addresses the diversity gap in the cognitive genetic research and presents an opportunity to improve the population health. Our study identifies opportunities to address both cardiometabolic and cognitive health at population levels, maintaining human potentials and health in this ageing world.

## Methods

### Phenotypic and genomic data derivation

Participants’ in-depth phenotypes were derived from the HELIOS study (ethic approval: IRB-2016-11-030, www.healthforlife.sg), a baseline population cohort comprising the multi-ethnic Asian population of Singapore. The inclusion criteria were Singaporean citizens or Permanent Residents aged 30-84 years old. The exclusion criteria were pregnancy, breastfeeding, major illness/ surgery, or participation in drug trials within the past month, and cancer treatment in the past year. Participants were recruited from the general population to ensure diversity in ethnicity and socio-economic background. The three primary ethnic groups in Singapore are: i. Chinese; ii. Malay, and iii. Indian. All participants underwent standardized HELIOS study protocol^9^. Participants were asked to fast for ≥8 hours for all biological samples. All physiological assessments were compulsory, except when contraindications were present, or when participants experienced discomfort or fatigue such as during repeated measures.

Cognitive function was assessed with multi-domain computer-based test adopted from the UK Biobank, as previously described.^38^ The protocol details for all physiological assessments, laboratory assays and extensive health and lifestyle questionnaires are available in the cohort methodological paper^9^. The physical activity levels were further adjusted with 7-day accelerometry^9^. **S Method 1** described the calibration process of the physical activity levels, and **S Method 2** explained the derivation of food groups.

Whole genome sequencing was performed using the Novaseq platform and data processed using DRAGEN v3.7.8, followed by creation of HAIL matrix tables and downstream stringent QC, as previously described^9^. The QC steps for the population genetic structure analysis are also available in our methodological paper^9^.

### PheWAS (epidemiological) analysis

Analysis was completed in R version 4.4.0. We reviewed a catalogue of 3190 variables from the HELIOS study. After the removal of operational variables, we conducted triage based on the characteristics of research variables, research domains, and variable types, and identified 872 suitable variables (**S Fig 1**). We aggregated variables derived from repeated measures and calculated composite scores using individual or domain variables where appropriate (for example, Pittsburgh Sleep Quality Index), shortlisting a total of 542 variables suitable for downstream PheWAS (**S Fig 1**). We excluded participants who did not complete cognitive function test (**S Figure 1**). Excluded participants were older, more likely to be female, Indian, with lower education and have poorer cardiometabolic profile (**S Table 1**). We derived a single proxy of general cognition ‘***g***’ by dimensionally reducing z-normalized cognitive test raw scores using a confirmatory factor analysis^38^, as previously established in the GWAS of general cognition.^12^ All laboratory assay results were ln-transformed. All continuous variables were subsequently z-scored. Genetic Principal Components (PCs) were derived using Principal component analysis (PCA)^9^.

We then performed multiple linear regressions with ‘***g****’* as exposure variable, adjusting for sex, age, ethnicity, the interaction of sex and age, age-squared, sex and age-squared, and the first 5 genetic PCs. The resulting p values were adjusted for multiple comparisons using the Benjamini & Hochberg method to control for false discovery rate. We repeated the PheWAS with reaction time as alternative exposure variable as sensitivity analysis. We performed additional hierarchical clustering on the intra-correlations of phenotypes across under-represented assessment domains to further probe phenotypes with p at the marginal regions. Optimal cluster numbers were determined by average silhouette width (*factoextra* package) and heatmap visualisation (*ComplexHeatmap* package). We then selected phenotypes with the smallest p-value when multiple phenotypes emerged from a single cluster.

### Mendelian Randomization (MR)

To perform Two-sample MR, we shortlisted the equivalent phenotypes in the UK Biobank, in which the Genome-Wide Association Studies (GWAS) summary statistics are available in the IEU Open GWAS project (https://gwas.mrcieu.ac.uk/). We deliberately restricted the search to statistics derived from male and female Europeans to minimize potential heterogeneity in the summary statistics. We prioritised statistics with the largest sample size and the latest publication year. We selected statistics generated using Phesant pipeline from the UK Biobank49, but would select those with larger sample size from international consortia if available.

Two-sample MR was performed to obtain causal evidence linking ‘***g***’ and multi-domain phenotypes using *TwoSampleMR* R package. *F*-statistics per instruments were calculated to assess weak instrument bias50. The default *TwoSampleMR* clumping procedure was applied to the instruments. The Single Nucleotide Polymorphism (SNP) from both instrument-exposure and outcome were then harmonized to the forward strand, and any ambiguous SNPs were excluded. We performed inverse-variance-weighted MR and weighted-median MR, and applied Bonferroni significance threshold for 0.05/23= P <0.002 to account for multiple testing. To test for horizontal pleiotropy, we performed MR-Egger, leave-one-out, single-SNP MR analysis, and repeated the MR after SNPs predicting alternative phenotypes (verified against the GWAS catalog using *gwasrapidd*) were removed. For repeat Two-sample MR without pleiotropic SNPs, we maintained SNPs causally linked with *brain region volumes* and any phenotypes that contain the following elements: *cognition, cognitive function, creativity, education, reading, reasoning, executive function, and math ability*. We performed MR with ‘***g***’ as instrument-outcome to evaluate reverse causality.

### Functional annotation

After removing pleiotropic SNPs, we extracted the Reference SNP-cluster IDentification (rsID) of 134 SNPs and performed functional annotation using the web interface of Variance Effect Predictor in Ensembl (https://grch37.ensembl.org/Homo_sapiens/Tools/VEP). To ascertain tissue specificity of candidate genes, we aggregated all SNPs predicting ‘***g***’ from the MR that demonstrated causality, resulting in 144 genes after removing redundancies. We then submitted the candidate gene symbols of (Functional Mapping and Annotation) FUMA platform51 (https://fuma.ctglab.nl/) and selected GTEx v8 54 tissue types52. To further confirm broad representation of gene functions, we performed enrichment analysis using *clusterProfiler53* with cut-off p=0.05.

### Comparison of allele frequencies across ancestries

Using a similar rsID list of 134 SNPs, we extracted minor allele frequencies across 8 different ancestries from Allele Frequency Aggregator or ALFA, a feature in dbSNP (https://www.ncbi.nlm.nih.gov/snp/docs/gsr/alfa/). The 8 ancestries include African, Asian, European, Latin America 2, Latin America 1, Other, and South Asian and Total (multi-ancestry). To determine if the allele frequencies were significantly different across ancestral groups, we performed ANOVA test and aggregated the allele frequency into decile per ancestries for plotting.

## Data availability

Data access request can be submitted to the HELIOS Data Access Committee by emailing for details. Dummy data and latest version of data dictionary can be found in https://www.npm.sg/research/call-for-proposals/.

## Acknowledgements

T.M. is funded by the Lee Kong Chian School of Medicine Dean’s Postdoctoral Fellowship. J.C. is supported by Singapore Ministry of Health’s (MOH) National Medical Research Council (NMRC) under its OF-LCG funding scheme (MOH-000271-00), Singapore Translational Research (StaR) funding scheme (NMRC/StaR/0028/2017), the National Research Foundation, Singapore through the Singapore MOH NMRC and the Precision Health Research, Singapore (PRECISE) under the National Precision Medicine programme (NMRC/PRECISE/2020) and intramural funding from Nanyang Technological University, Lee Kong Chian School of Medicine and the National Healthcare Group. M. Lam is supported by the Singapore Ministry of Health’s National Medical Research Council under its Centre Grant (CG21APR1002). We thank Matthias Liau Yi Quan for the initial factor analysis for ***g***.

## Author contributions

M.Lam, J.C., J.L., T.M. conceived and designed the investigation. T.M., and the HELIOS Study team implemented the study and collected data. T.M., N.S, D.L.Y.W., P.R.J, X.W., H.K.N curated epidemiological data. T.M. performed the data analyses. M.Lam. supervised the analyses. T.M and M.Lam wrote the initial draft manuscript. All authors reviewed and contributed to the revision of the submitted manuscript.

## Code availability

The analytic codes will be made available via GitHub repository https://github.com/HELIOS-SG100K-LKC/PheWAS-cognition.git.

## Supplementary Note

### The HELIOS Study team comprises

John C Chambers^1,4,11^, Marie Loh^1,10^, Paul Elliott^1,4^, Lee Eng Sing^1,6^, Jimmy Lee^1,2^, Joanne Ngeow^1,5^, Sabrina Wong^1,7^, Elio Riboli^1,4^, Tricia Chang^7^, Rinkoo Dalan^1,12^, Wai Kee Kok^7^, Benjamin Lam Chih Chiang^1,13^, Kelvin Li1^1,14^, Lim Tock Han^1,14^, Pritesh R Jain^1^, Hong Kiat Ng^1^, Theresia Mina^1^, Nilanjana Sadhu^1^, Akash Bahai^1^, Dorrain Yanwen Low^1^, Xiaoyan Wang^1^, Harinakshi Sanikini^1,4^, Darwin Tay^1^, Terry Tong Yoke Yin^1^, Kostas Tsilidis^1,4^, Gervais Wanseicheong^1,15^, Yew Yik Weng^1,1^

The affiliations of the main co-authors and the HELIOS Study team are:

1 Lee Kong Chian School of Medicine, Nanyang Technological University, Singapore

2 Research Division, Institute of Mental Health, Singapore

3 Genome Institute of Singapore, Agency for Science, Technology and Research, Singapore

4 Department of Epidemiology and Biostatistics, School of Public Health, Imperial College London, United Kingdom

5 Division of Medical Oncology, National Cancer Centre, Singapore

6 MOH Office for Healthcare Transformation (MOHT)

7 National Healthcare Group Polyclinics, Singapore

8 Department of Neurology, National Neuroscience Institute, Singapore

9 Duke-NUS Medical School, Singapore

10 National Skin Centre, Research Division, Singapore

11 Precision Health Research (PRECISE), Singapore

12 Department of Endocrinology, Tan Tock Seng Hospital, Singapore

13 Khoo Teck Puat Hospital, Singapore

14 Department of Ophthalmology, Tan Tock Seng Hospital, Singapore

15 Department of Diagnostic Radiology, Tan Tock Seng Hospital, Singapore

## Supplementary Methods

### 1. Deriving equations to adjust iPAQ-derived MET levels

We processed the accelerometry raw data of a subset comprising 647 participants with Wave (https://github.com/MRC-Epid/Wave)^50,51^ to generate mean Euclidean Norm Minus One (ENMO). Briefly, Wave calibrated the data to local gravity based on stationary periods to reduce technical device-level measurement error. Next, noise was filtered out using a low pass Butterworth filter, with a frequency cut-off of 20Hz. Non-wear time was defined as consecutive stationary periods lasting for ≥1hr where all three acceleration axes had a standard deviation of <13.0 mg. ENMO across valid wear-time 5-seconds epoch was then calculated using 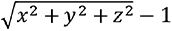, expressed in miligravity. A valid wear-time is defined as a minimum of 72-hour of wear and minimum 7 hours of wear in each “quadrant” of the day (12-6am, 6am-12pm, 12-6pm, 6pm-12am) over 7 days.

In the accelerometry subset, we defined ENMO∼ MET_iPAQ_*a + constant (*Equation 1*). We applied *Equation 1* to the full HELIOS dataset to predict ENMO. Since the variance of predicted ENMO (x) ≠ the variance of actual ENMO (x^*^),

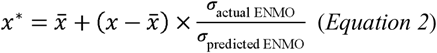

… where x = mean predicted ENMO and = standard deviation.

After applying *Equation 2* to *Equation 1*, the corrected MET_iPAQ_ could be calculated. Using the 647 accelerometry dataset, we obtained ENMO = 1.013247*MET_iPAQ_*10^-3^) + 25.14666. Due to the lack of Asian data, here we used Activity Energy Expenditure, or AEE= 5.01 + 1.000*ENMO, in J min-1 Kg-1 based on the UK study.^51^ Given that 1 MET = 4184 J kg^-1^ h^-1^ = (4184/60) J kg^-1^ min^-1^, 1 AEE = 60/4184 MET, and given that the number of minutes/ week = 10,080, the corrected MET_iPAQ_ in this study was:

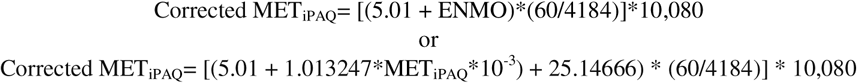

### 2. Calculation of dietary indices

Participants whose total energy intake <500 Kcal/day or > 3SD in men and women were excluded^52^. We also applied the Goldberg methods which considers individual physical activity levels as a sensitivity analysis^53^. Macronutrient information encompasses carbohydrates, proteins, starch, sugar, fibres, total fat, saturated, mono- unsaturated, and poly-unsaturated Fatty Acid (SFA, MUFA and PUFA, respectively), where carbohydrates are defined as the sum of starch and sugar58. Energy and macronutrients per food item = serving/day * weight/portion * macronutrients/serving and were subsequently aggregated to calculate total energy and macronutrients per day per individual. Macronutrients were expressed as the proportions of the estimated total daily energy intake by applying Food and Agricultural Organization-recommended energy density (proteins, carbohydrates, starch, and sugar = 4 *Kcal/g*, fats=9 *Kcal/g*, and fibre= 2 *Kcal/g*59). Food items were grouped as previously described^52^. After alignment, total energy and macronutrient intake per food group were derived.

**Supplementary Figure 1.**
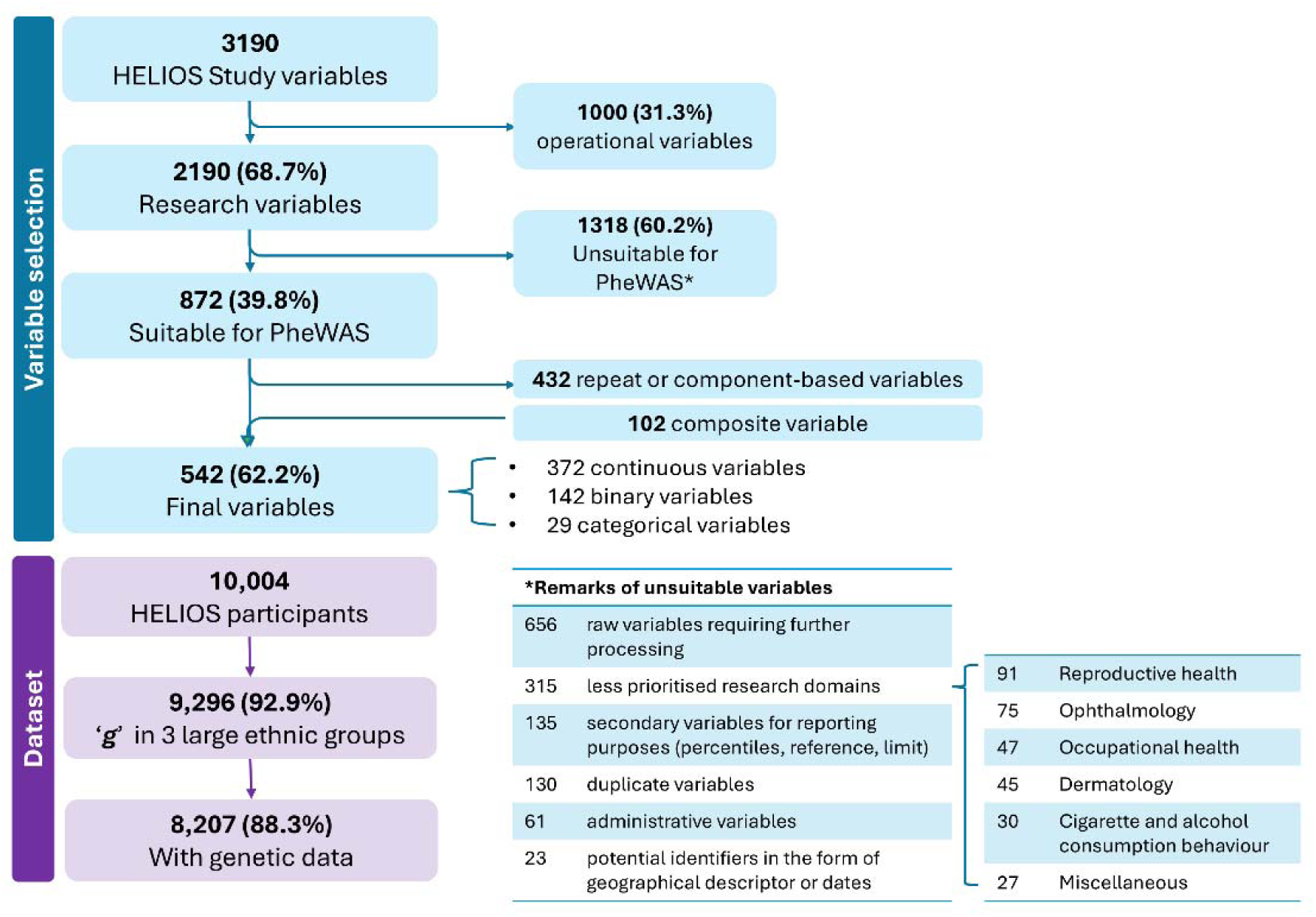
CONSORT diagram of the study. Some research domains were deprioritised due to the very small sample size following skip logic applications (E.g., “Number of cigarette smoked before quitting” was applicable to participants who have reported as smoker or ex-smoker, in addition the unique, low cigarette smoking prevalence in Singapore), or because data were not yet ready (E.g., retinal imaging).

**Supplementary Figure 2.**
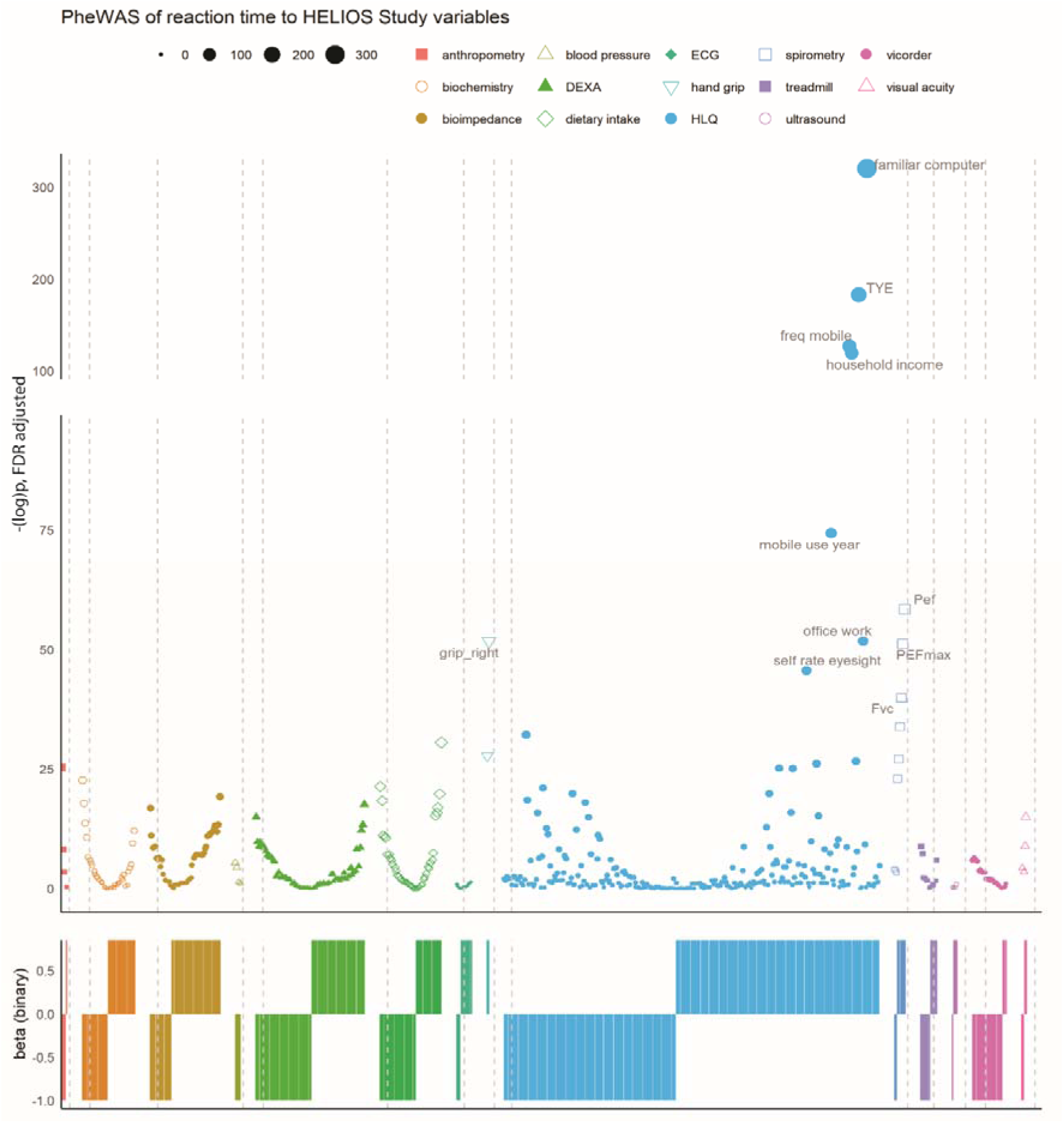
Phenome-wide Association Studies (PheWAS) of reaction time in multiethnic Asian populations. PheWAS was conducted with reaction time as exposure variable and 542 research variables as outcome variables, excluding ‘***g***’. The research variables were grouped according to the HELIOS Study assessments (x-axis), in alphabetical order of assessments. Colour and shape represent different assessment domains, separated by grey dotted lines. Circle size represents the magnitude of -log(P). All continuous variables were z normalised. All regressions were adjusted for sex, age, ethnicity, interaction effect of age-squared, and interaction effect of gender and age-squared. The variable coding was such that higher scores indicate greater values of outcome variables, unless otherwise indicated. To illustrate the direction of the associations, β was converted to binary outcome. *DEXA*: Dual X-Ray Absorptiometry; *ECG*: electrocardiogram; *TYE*: total year of education; *freq mobile*: frequency of mobile phone usage/week; *FEV1*: Forced Expiratory Volume; *FVC*: Forced Vital Capacity; *HLQ*: health and lifestyle questionnaire; *PEF*: Peak Expiratory Flow.

**Supplementary Figure 3.**
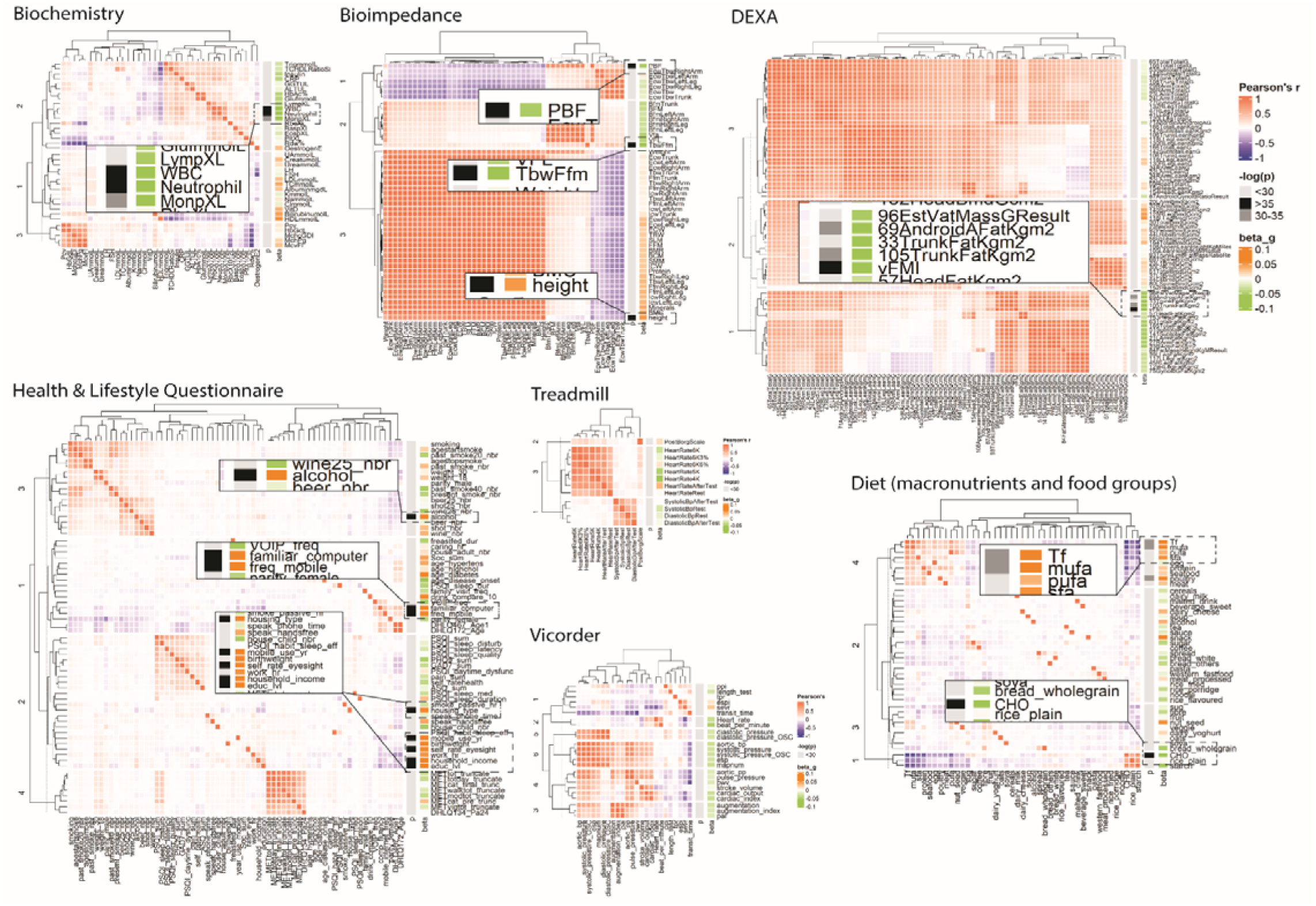
The hierarchical clustering of correlation matrices to determine additional variables for two-sample MR of ‘*g*’ with phenome, to identify additional variables at borderline (depicted in dark grey, -log(p)=30-35) which did not cluster with variables identified in the main PheWAS plot (black).

**Supplementary Figure 4.**
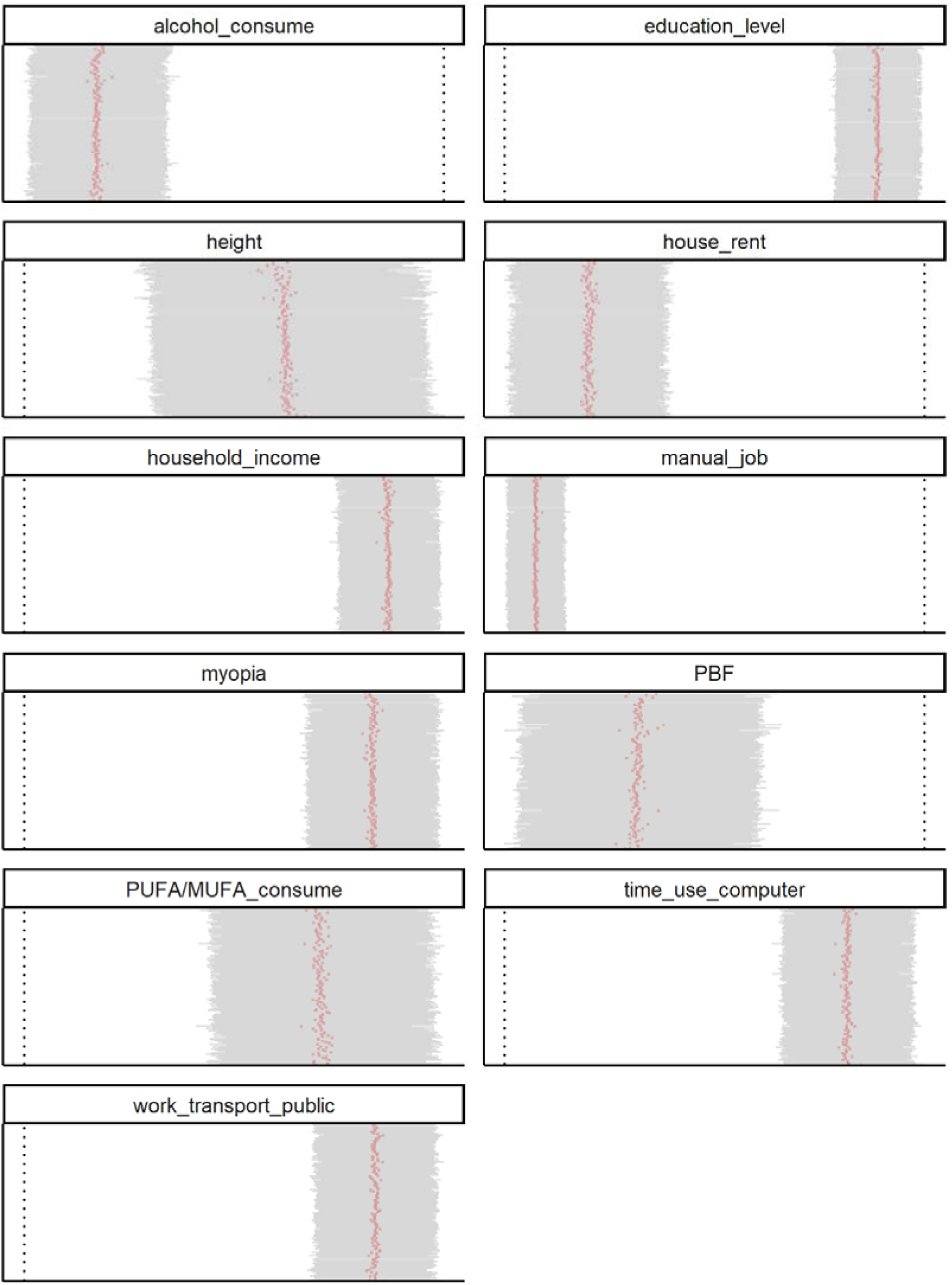
The leave-one-out MR for 12 phenotypes with genetically associated with ‘*g*’. The dotted line is OR=1. Each red dot is 1 leave-one-out MR with ‘***g***’ as instrument-exposure. and grey lines are 95% CI. *MUFA*: mono-unsaturated fatty acids; *PBF*: Percent Body Water; *PUFA*: poly-unsaturated fatty acids.

**Supplementary Figure 5.**
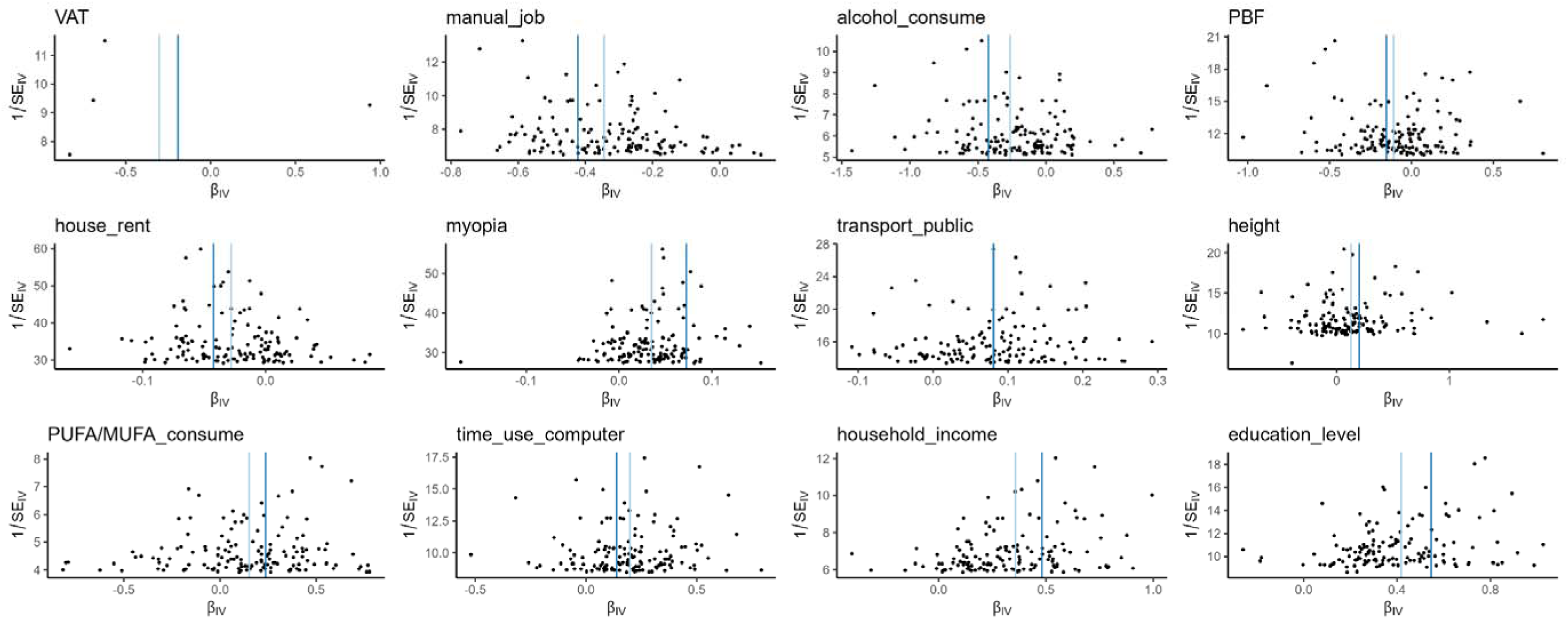
Funnel plots depicting Single-SNP MR. Out of 8 SNPs in harmonised data with VAT as instrument-outcome, 4 were excluded. *MUFA*: mono- unsaturated fatty acids; *PBF*: Percent Body Water; *PUFA*: poly-unsaturated fatty acids.

**Supplementary Figure 6.**
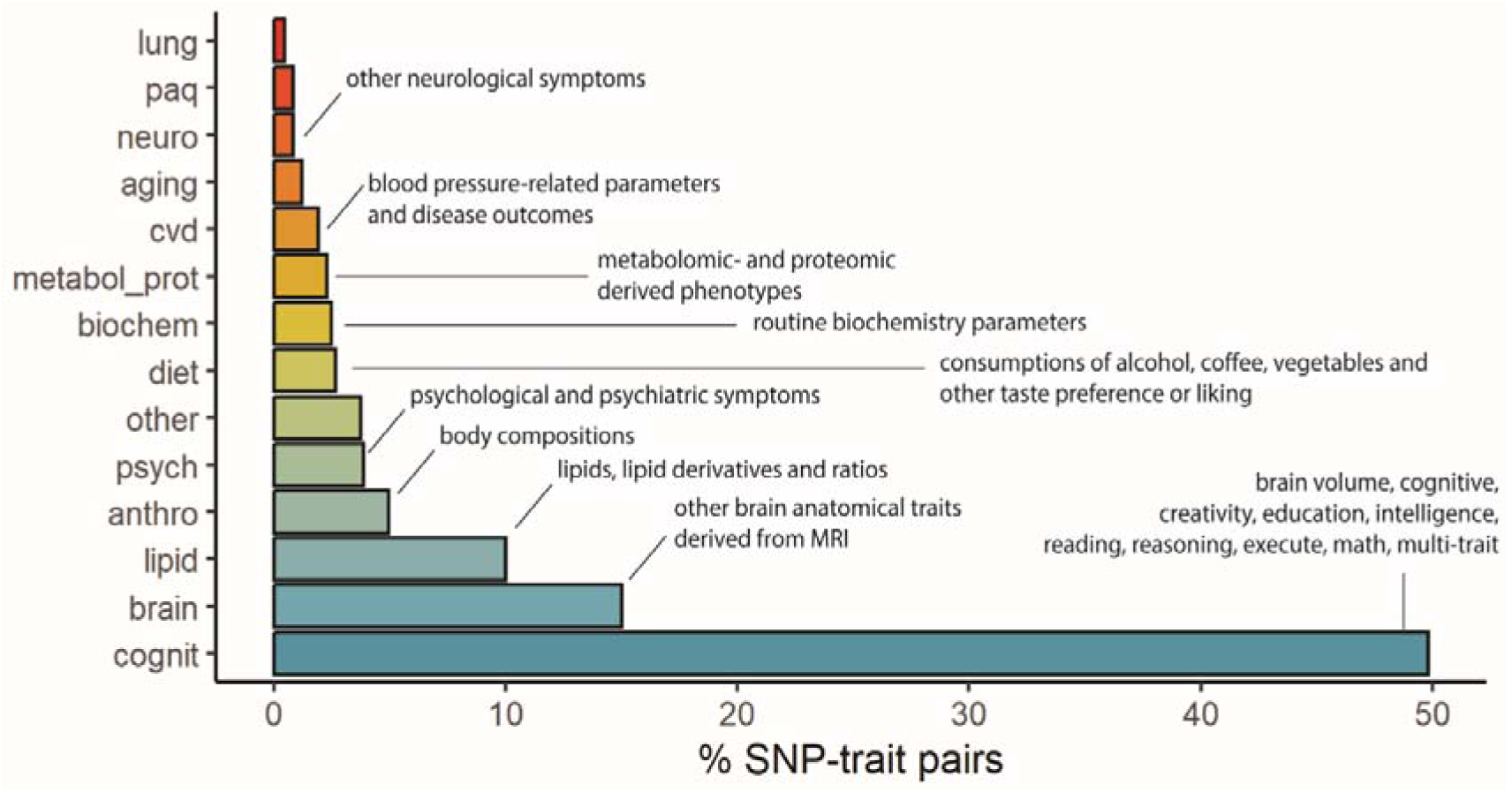
Breakdown of pleiotropic SNPs. There are 1062 SNP-phenotype combinations available in the GWAS catalogue from the summary statistics of ‘***g***’. Domain description is provided in the figure, for further details of the classification, see **Supplementary** Table 8. *Aging*= aging-related phenotypes such as male balding patterns, grip strength, age of menarche; *lung*= lung function; *paq*=physical activity; *other*= Other phenotypes such as sociodemographic parameters, personality traits, and non-cardiovascular disease outcomes.

**Supplementary Figure 7.**
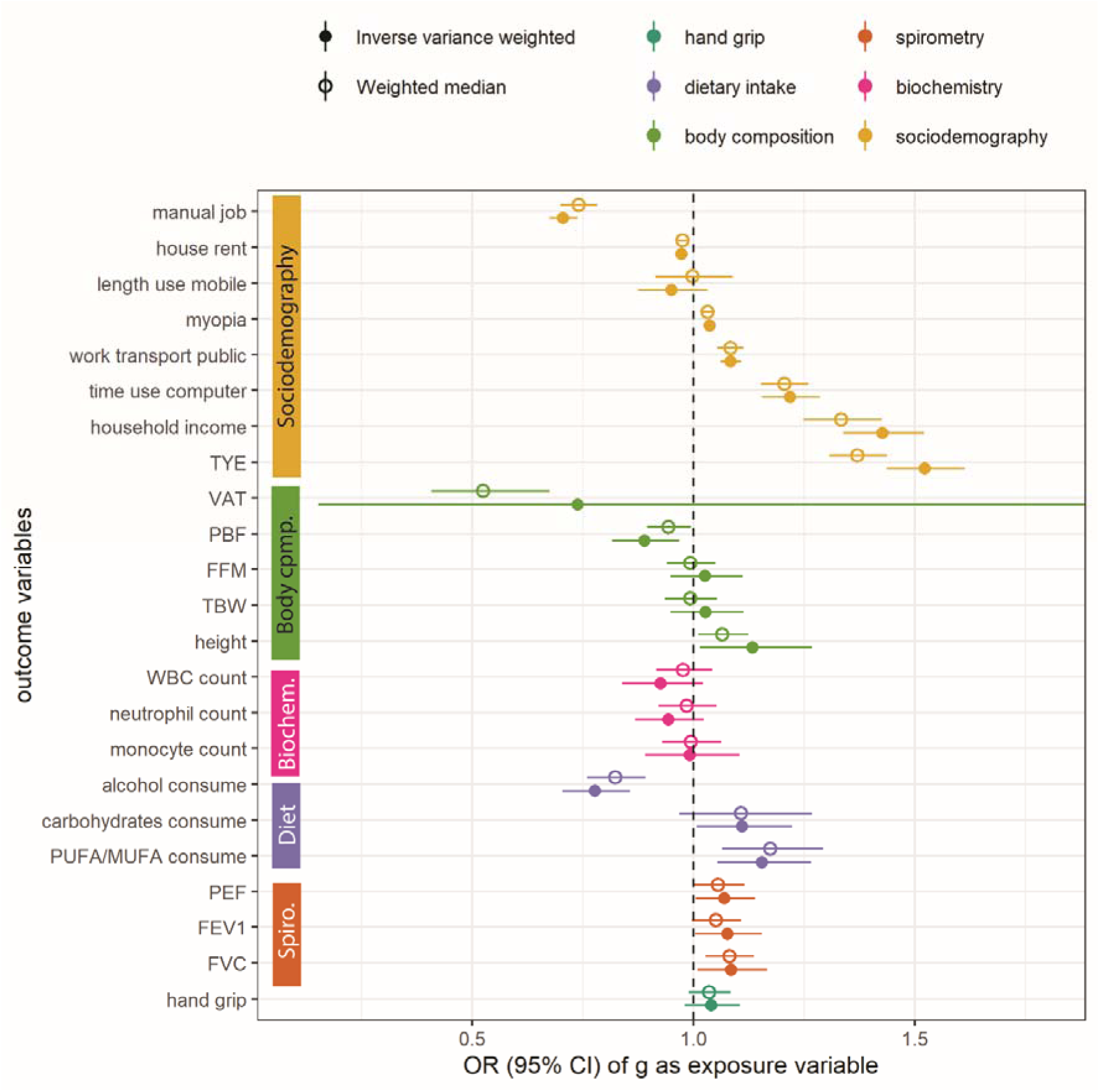
Two-sample Mendelian Randomisation (MR) of general cognitive function ‘*g*’ without pleiotropic SNPs. The ‘*g*’ was instrument-exposure. *FEV1*: Forced Expiratory Volume; *FFM*: fat-free mass; *FVC*: Forced Vital Capacity; *MUFA*: mono-unsaturated fatty acids; *PBF*: Percent Body Water; *PEF*: Peak Expiratory Flow; *PUFA*: poly-unsaturated fatty acids; *TBW*: total body water; *TYE*: total year of education; *VAT*: DEXA-based visceral fat mass; *WBC*: white blood cell.

**Supplementary Figure 8.**
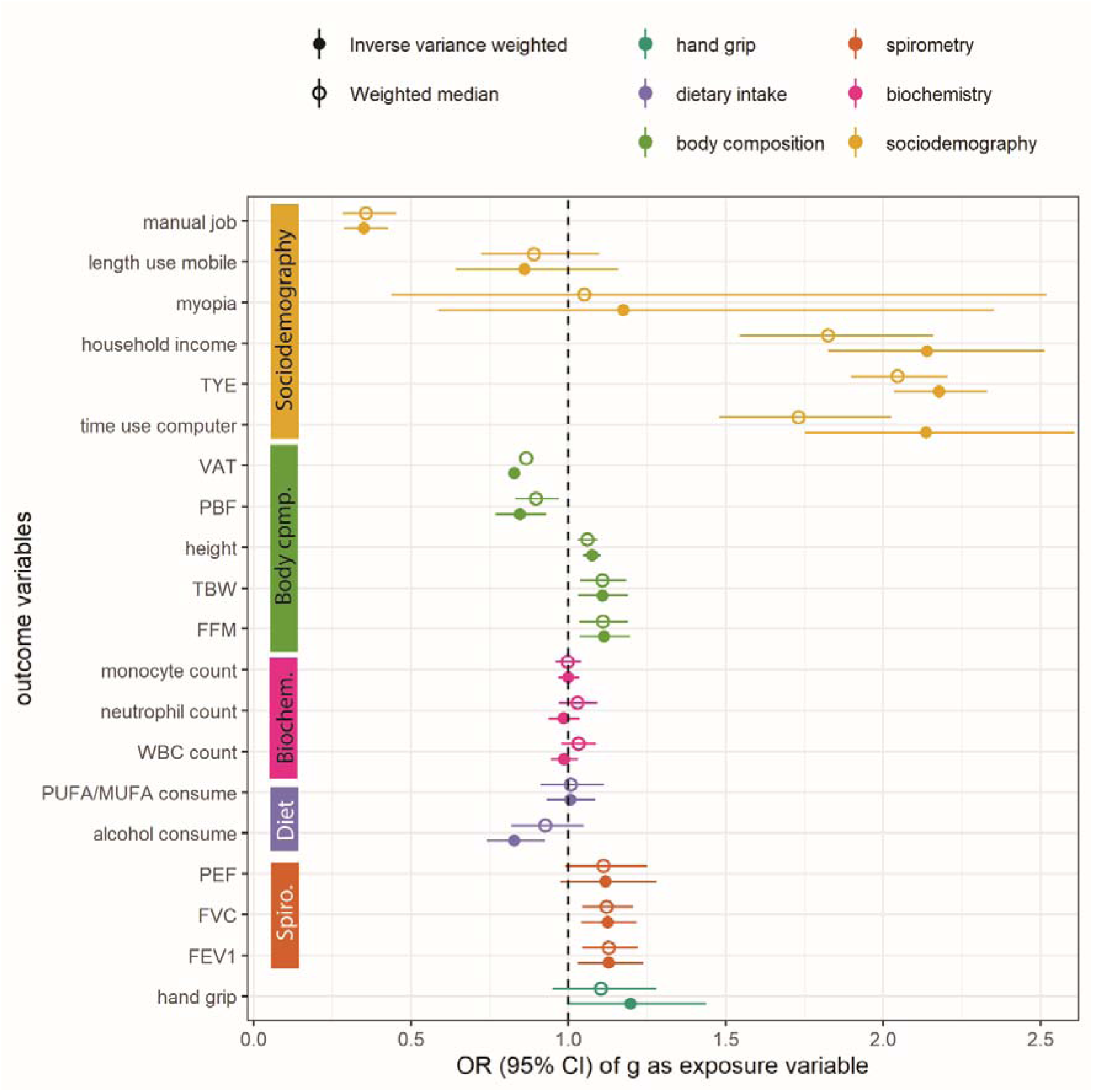
Reverse Two-sample Mendelian Randomisation (MR) of general cognitive function ‘*g*’. Carbohydrate, use of public transport and house renting were removed due to insufficient SNPs to conduct MR. The ‘***g***’ was instrument-outcome. *FEV1*: Forced Expiratory Volume; *FFM*: fat-free mass; *FVC*: Forced Vital Capacity; *MUFA*: mono-unsaturated fatty acids; *PBF*: Percent Body Water; *PEF*: Peak Expiratory Flow; *PUFA*: poly-unsaturated fatty acids; *TBW*: total body water; *TYE*: total year of education; *VAT*: DEXA-based visceral fat mass index; *WBC*: white blood cell.

**Supplementary Figure 9.**
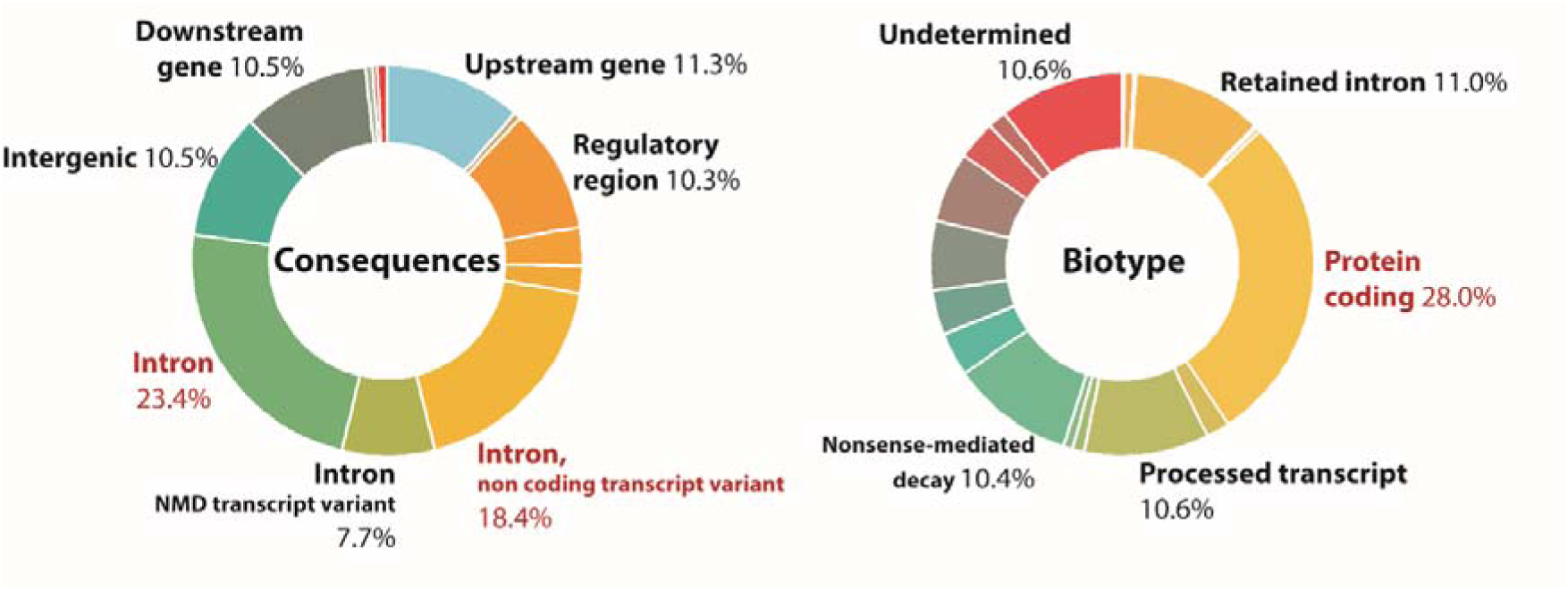
Summary statistics of variant effect predictors for SNPs predicting general cognitive function ‘*g*’.

**Supplementary Figure 10.**
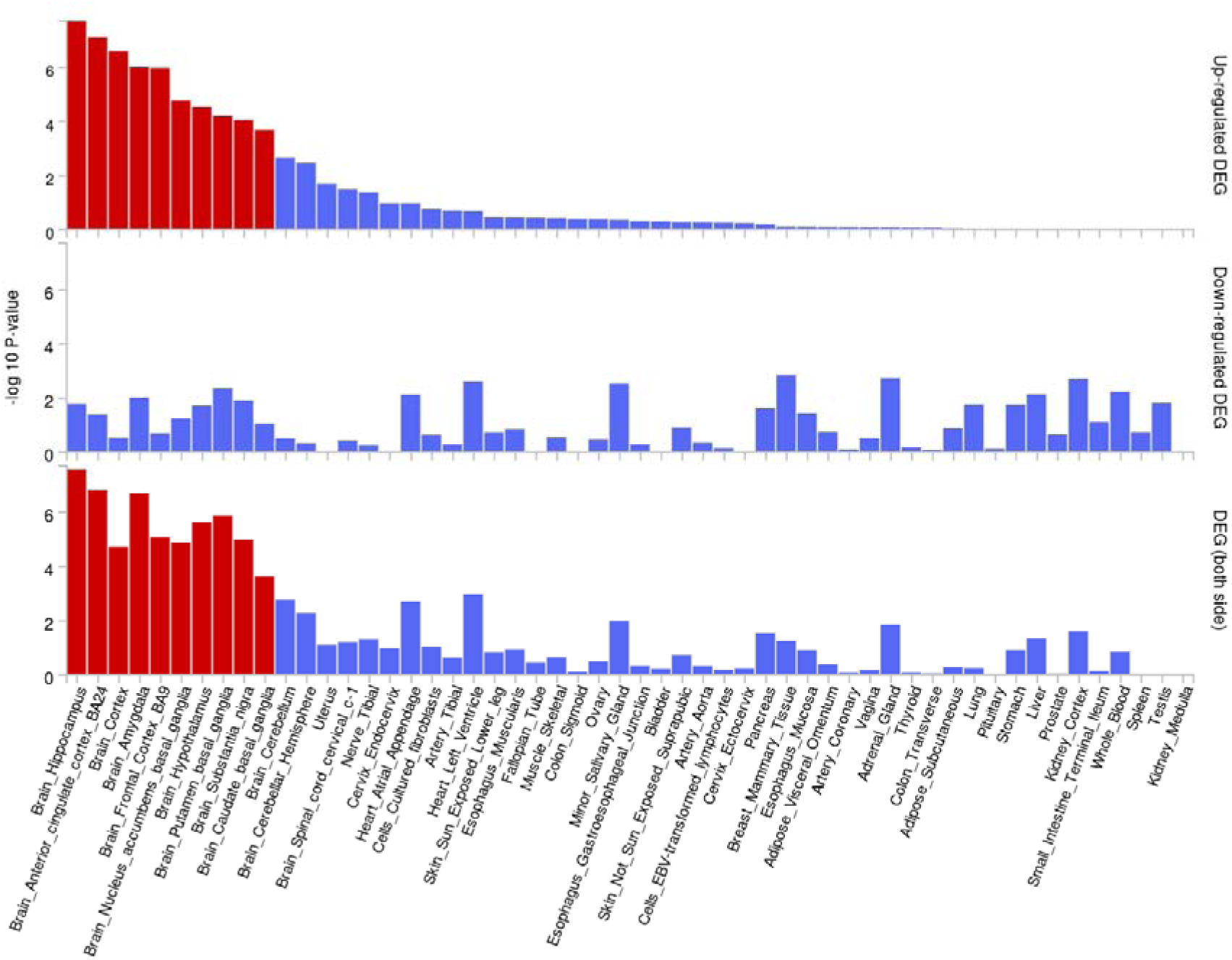
Candidate genes linked general cognitive function ‘*g*’ are over-expressed in brain sub-tissues. There were 144 genes linked to 134 SNPs predicting general cognitive function ‘*g*’. The barplot was generated with FUMA platform51, using GTEx v8 54 tissue types52.

**Supplementary Figure 11.**
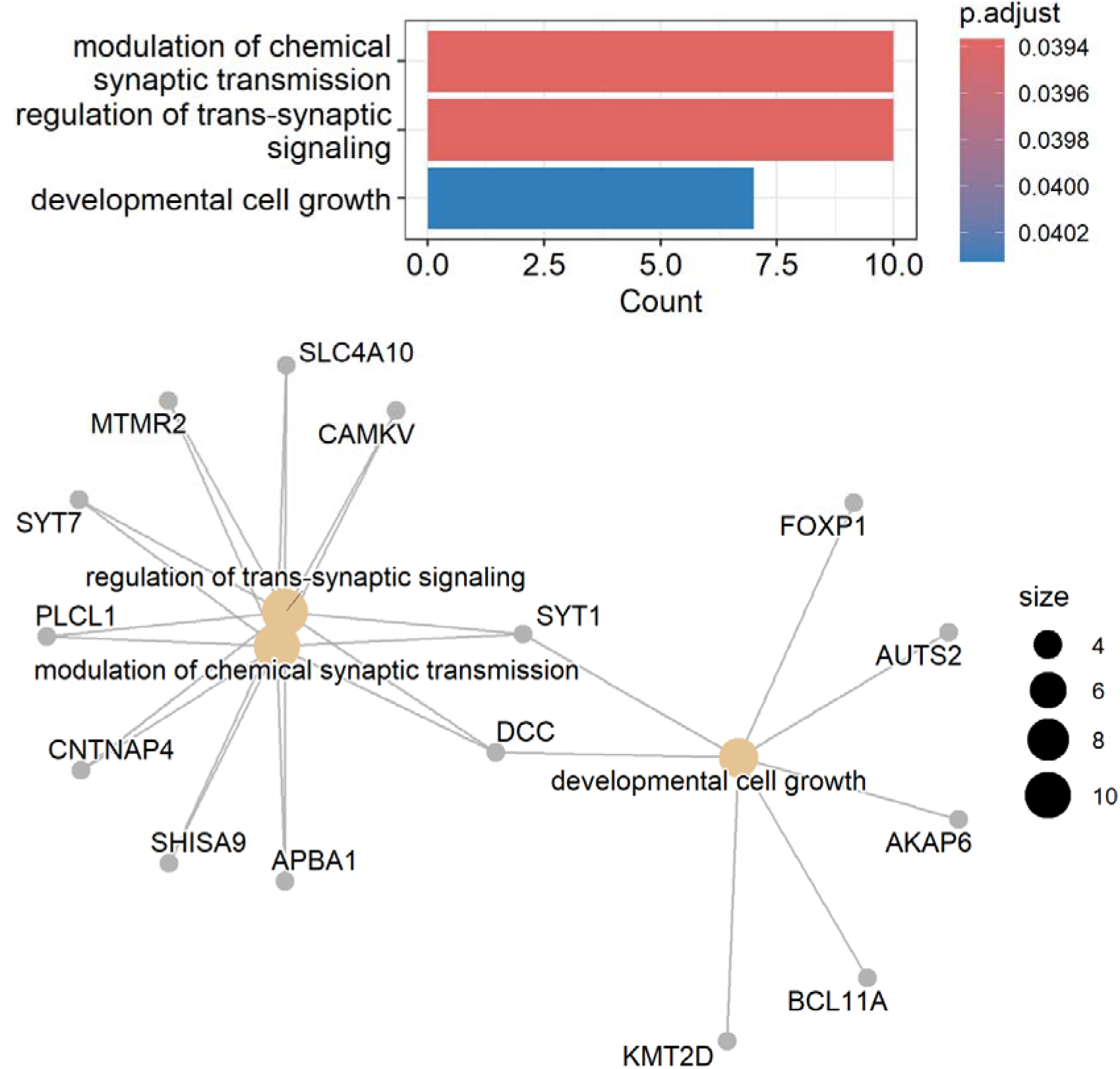
**Broad representation of gene functions linked to general cognitive function ‘*g*’**. Enrichment analysis was performed on 106 out of 144 candidate genes with cut-off p=0.05.

**Supplementary Figure 12.**
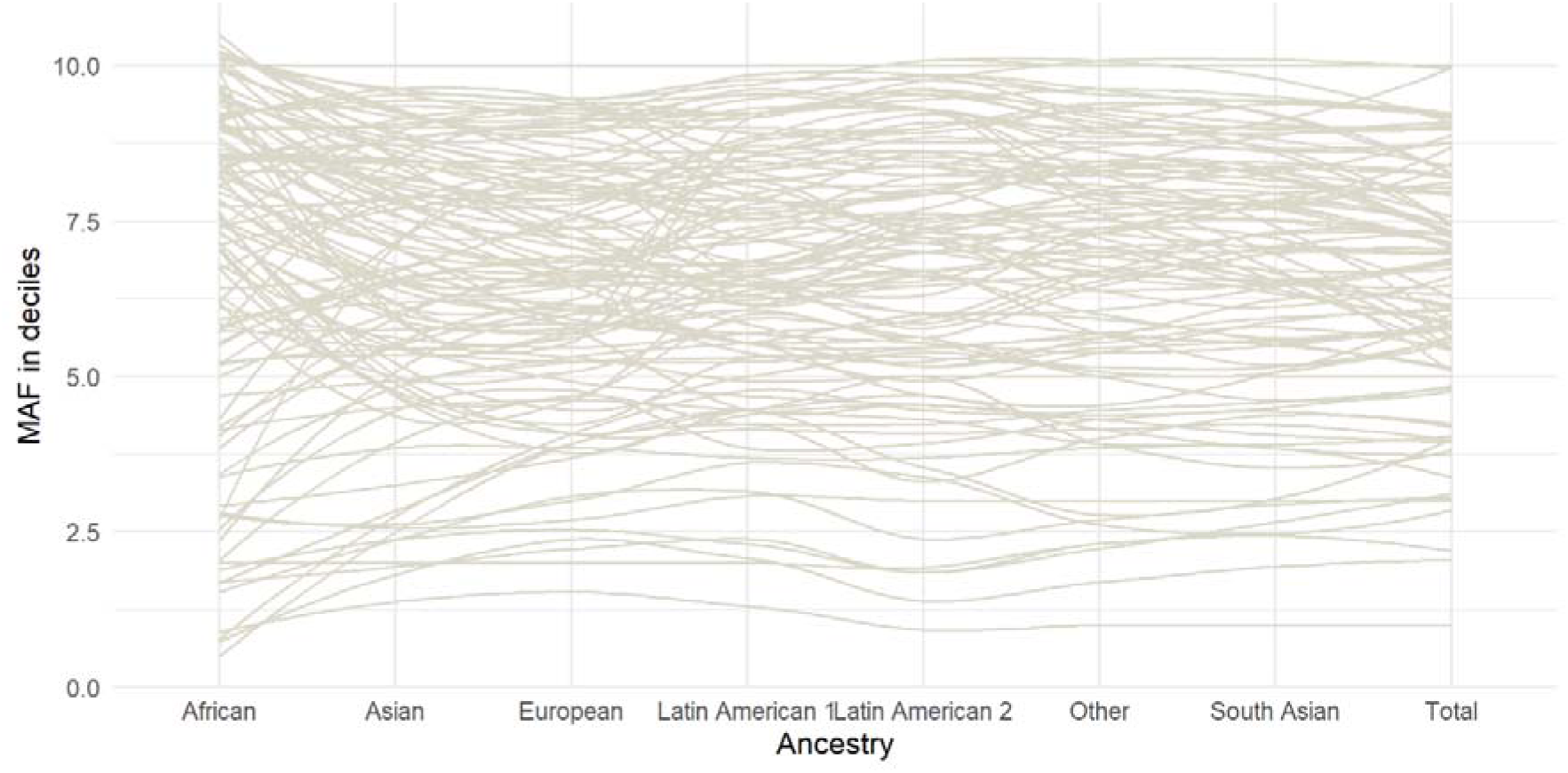
Minor allele frequency comparison of SNPs predicting general cognitive function ‘*g*’. There are 134 unique SNPs. P_ANOVA_ across 8 ancestries was p=0.243. We extracted minor allele frequencies across 8 different ancestries from Allele Frequency Aggregator (https://www.ncbi.nlm.nih.gov/snp/docs/gsr/alfa/).

